# ML for MI - Integrating Multimodal Information in Machine Learning for Predicting Acute Myocardial Infarction

**DOI:** 10.1101/2022.10.25.22281536

**Authors:** Ran Xiao, Cheng Ding, Xiao Hu, Jessica Zègre-Hemsey

## Abstract

Early identification and recognization of myocardial ischemia/infarction (MI) is the most important goal in the management of acute coronary syndrome (ACS). The 12-lead electrocardiogram (ECG) is widely used as the initial screening test for patients with chest pain but its diagnostic accuracy remains limited. There is an ongoing effort to address the issue with machine learning (ML) algorithms which have demonstrated improved performance. Most studies are designed to classify MI from healthy controls and thus are limited due to the lack of consideration of potential confounding conditions in the setting of MI. Moreover, other clinical information in addition to ECG has not yet been well leveraged in existing machine learning models. The present study aims to advance ML-based prediction models closer to clinical applications for early MI detection. The study considered downstream clinical implementation scenarios in the initial model design by dichotomizing study samples into MI and non-MI classes. Two separate experiments were then conducted to systematically investigate the impact of two important factors entrained in the modeling process, including the duration of ECG (2.5s vs. 10s), and the value of multimodal information for model training. A novel feature-fusion deep learning architecture was proposed to learn joint features from both ECG and patient demographics as the additional data modality. The best-performing model achieved a mean area under the receiver operating characteristic curve (AUROC) of 92.1% and a mean accuracy of 87.4%, which is on par with existing studies despite the increased task difficulty due to the new class design. The results also show that the ML model can capitalize on the information added from both the extra ECG waveforms in time and patient demographics. The findings in this study help guide the development of machine learning solutions for early MI detection and move the models one step closer to real-world clinical applications.

## Introduction

Cardiovascular disease (CVD) remains the leading cause of death in the United States and around the world. Acute coronary syndrome (ACS) is the acute manifestation of myocardial ischemia/infarction (MI) and relies on rapid and accurate identification. Although nearly 6 million adults in the US are evaluated annually for chest pain; among those evaluated only a small subset are diagnosed with ACS^1, 2^. The most important goal in the management of ACS is identifying the problem early and initiating prompt intervention, which is based on the premise that longer delays are associated with worse clinical outcomes^3^.

To date, the 12-lead electrocardiogram (ECG) remains the most widely useful initial screening test for evaluating patients with chest pain. Considered the “gold standard” for non-invasive electrocardiographic detection of acute myocardial ischemia/injury, the standard 12-lead is noninvasive, inexpensive, and the single most important method for rapid detection of ACS in emergency care settings (i.e., prehospital, emergency department)^4^. Current American Heart Association (AHA) guidelines recommend ECG acquisition within 10 minutes of first medical contact for individuals with symptoms suggestive of ACS including, but not limited to, non-traumatic chest pain, shortness of breath, radiating pain to arm or jaw, nausea, diaphoresis, or lightheadedness. The diagnostic accuracy of ECG for ACS by current threshold-based clinical rules, however, is limited (30-70% sensitivity, 70-95% specificity) ^5, 6^. Approximately 50%-55% of patients with acute myocardial ischemia and 40% with unstable angina do not have ST-segment elevation or depression present in their initial “snap-shot” ten-second 12-lead ECG recording^7, 8^. Continuous ECG monitoring, moreover, has shown many ischemic events are clinically silent and transient and thus may not be captured by a standard 12-lead ECG^9^. Another challenge to current ECG is the visual interpretation of ST-segment deviation, which is limited by high inter-rater variability, extensive time for analysis, and failure to detect subtle ischemic ECG changes. Each threatens the reliability and validity of the ECG.

Machine learning (ML) is data-driven and thus not limited by preset rigid features and thresholds inherent to the clinical rule-based algorithms, which has the potential to improve the diagnostic accuracy of MI. Many studies have recognized the opportunity and pioneered the implementation of machine learning for predicting MI. Wang et al. combined statistical and entropy features extracted from ECG waveforms and implemented principal component analysis (PCA) to reduce the dimension of features for training various shallow machine learning models to classify between MI and health control^10^. Similarly, Chang et al. extracted sequential features in the temporal dynamics of ECG using a hidden Markov model (HMM) to train a support vector machine (SVM) for the classification of MI and healthy control^11^. In another study, Sharma et al. utilized multiscale wavelet coefficients as features for detecting MI^12^. It is worth noting that the latter two studies do not consider patient assignments in the model development process (i.e., data from the same patient may exist in both training and test sets), which may artificially increase the model performance.

Inspired by promising achievements in the classification of cardiac arrhythmia, deep learning has gradually found its footing in studies for detecting MI as well. Reasat and Shahnaz used a convolutional neural network (CNN) and waveforms from three (i.e., lead II, III, and aVF) of the standard 12-lead ECG to classify between inferior myocardial infarction and healthy control^13^. Strodthoff et al. conducted a comprehensive study comparing the performance of seven different deep-learning architectures in classifying five diagnostic classes available in the PTB-XL dataset^14, 15^.

While the above studies set the stage for leveraging machine learning to improve diagnostic accuracy for MI using ECG, there are several limitations to current knowledge. First, most of the existing studies formulate the classification problem as MI vs. healthy control, which limits the clinical applications because many non-MI conditions can be present aside from the normal condition. Some of these conditions display electrocardiographic features confounding MI, such as left ventricular hypertrophy (LVH) and left bundle branch block (LBBB). Therefore, clinically viable algorithms should factor in these conditions. Second, existing studies use ECG of default length provided in the development dataset. One unanswered question is whether it suffices to use 2.5s of ECG data as it is presented to clinicians in a standard 12-lead ECG for training machine learning models. Using shorter data can improve computing efficiency whereas its impact on model performance is unknown. Third, other types of information routinely collected (such as patient demographics, past diagnoses, and symptoms) during the initial evaluation of MI have not been well incorporated into the machine-learning modeling process yet, despite existing evidence showing their diagnostic values in the initial evaluation phase for MI^16^.

The present study builds upon existing works^10-15^ and aims to advance machine learning-based prediction models closer to clinical applications for the early detection of MI. This was achieved by considering downstream clinical implementation in our experimental design and dichotomizing samples in the development dataset into MI and non-MI classes. We then conducted two separate experiments to systematically investigate the impact of two critical factors in the modeling process, the duration of ECG and the value of multimodal information entering model training. The purpose of this study was to use a novel feature-fusion deep learning architecture that can learn joint features from both ECG and patient demographics. The goal is to guide further work toward clinically oriented MI prediction models with improved performance.

## Methods

### Study data

We used a de-identified clinical ECG database for our study, the PTB-XL dataset, which is publicly accessible on the PhysioNet ^15, 17^. The dataset includes 21,837 clinical 12-lead ECG records in the duration of 10s. The data were collected from 18,885 patients who are 52% male (48% female), with a median age of 62 and an interquartile range of 22. Baseline patient characteristics (i.e., age and sex) and clinical annotations are provided along with each ECG record. At least one cardiologist (up to two) annotated ECGs with three diagnostic categories, i.e., statement, form, and rhythm, in accordance with the SCP-ECG standard^18^. Our study dichotomized ECG samples into two classes, a MI class (5,486 samples) and a non-MI class (16,351 samples). The non-MI class consists of samples that are annotated as either normal ECG (NORM), or as any of the following three diagnostic statements, ST/T change (STTC), Hypertrophy (HPY), and conduction disturbance (CD). The detailed breakdown of samples across different diagnostic statements can be found in Fig. 1.

**Figure 1.**
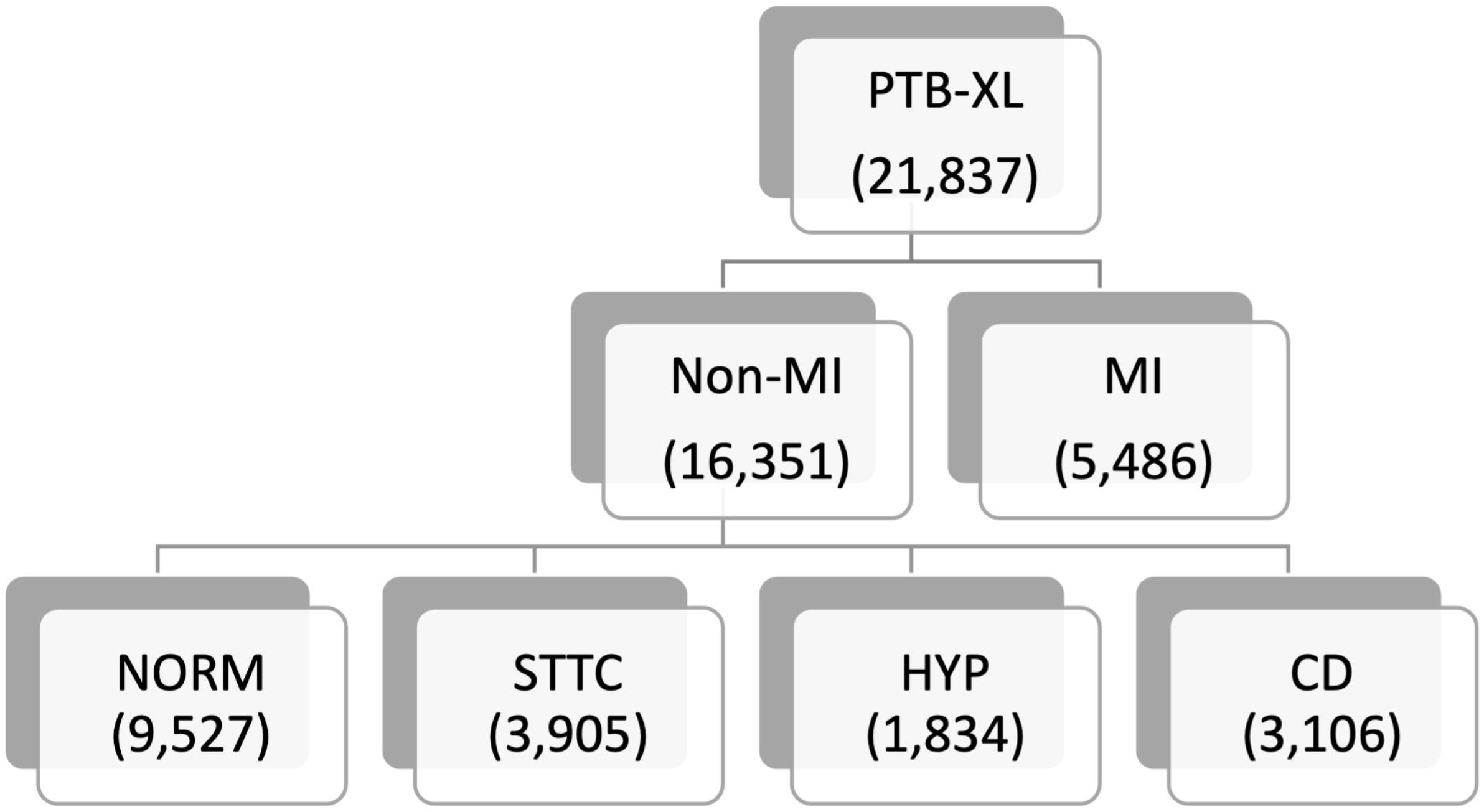
Breakdown of numbers of ECG samples across different diagnostic statements.

The dataset was prepared following a nine-fold cross-validation (CV) scheme, by first dividing it into ten equal partitions stratified across classes as provided by the PTB-XL dataset^15^. The tenth fold served as the independent test set. The remaining nine folders were cycled nine times. Within each cycle, one different fold of data served as the validation set, and the remaining eight folds served as the training data. The data split considers underlying patient assignments, by assigning samples of the same patient to the same partition. This avoids any data leakage between training and test sets, which could artificially boost model performance if not considered. The Institutional Review Board (IRB) of the local institute approved the study.

### Model development

We adopted two deep learning architectures that use different sources of inputs for the prediction of myocardial infarction. The ECG-only model used only ECG data for the model input. The 12-lead ECG waveforms were represented as 12 channels of one-dimensional temporal signals to train a 1D convolutional neural network, xResNet^19^. xResNet is a variant of the ResNet deep learning model^20^, by building upon the ResNet architecture and implementing a collection of practical structural tweaks that were proven to improve the performance over the vanilla ResNet model. xResNet was selected in the study as it is one of the top-performing models among various deep learning models tested on the PTB-XL dataset^14^. As shown in the yellow region of Fig. 2, 12-lead ECGs in the training set are input into the 1D xResNet model, which utilizes 101 convolutional layers to learn a comprehensive representation of ECG temporal dynamics, and outputs a prediction on the diagnosis of myocardial infarction.

**Figure 2.**
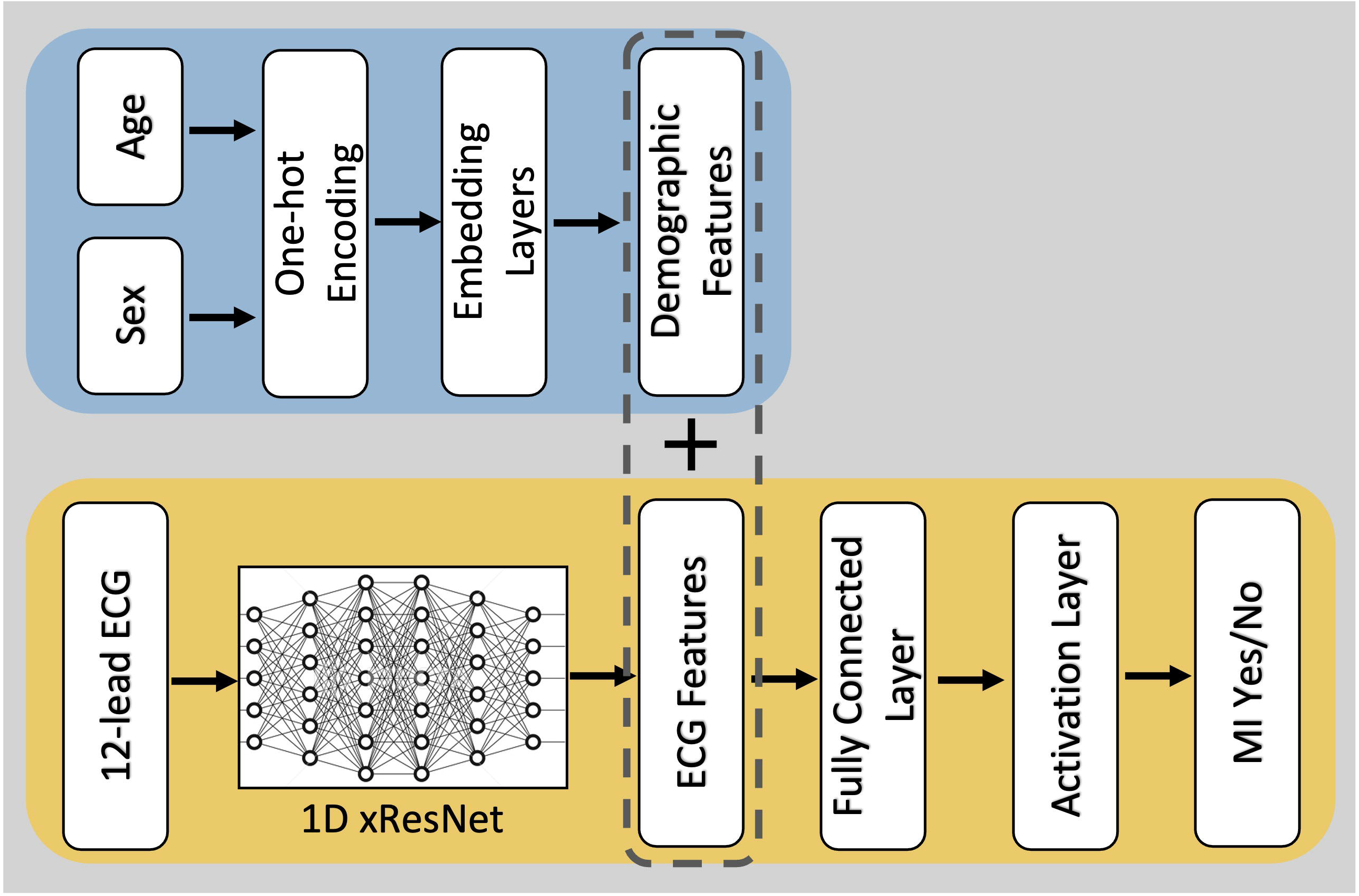
Schematic diagram of the ECG-only model (yellow) and the Multimodal model (yellow + blue).

The second model architecture bears our innovation in that it possesses a dedicated learner for patient demographic information (the blue region in Fig. 2), and integrates it with ECG features learned from the xResNet model (the yellow region in Fig.2) to form the final Multimodal model. The demographic learner consisted of three main components. The first component used one-hot encoding to achieve a numerical representation of different demographic variables. For age and sex, binary vectors with lengths of 99 and 2 were used to represent the corresponding patient information. Next, an embedding layer was deployed to encode patient demographics into a continuous vector space that could enhance the representation of underlying information. Age and sex information were embedded in vectors with lengths of 16 and 128, respectively. Lastly, both age and sex were upscaled to 512-element vectors through 1D convolutional layers, which generated the final demographic features. These features were concatenated (see region enclosed in the dashed box in Fig. 2) with the ECG features learned with the xResNet sans the last convolutional layer. The joint features were then input into a fully connected layer and an activation layer with sigmoid activation to yield prediction probabilities of MI.

### Training process

For each deep learning model and each fold of cross-validations, samples in the training set were cycled through 50 times (i.e., 50 epochs) with a batch size of 128. The final model was determined by comparing the validation loss from all training epochs and selecting the one that offered the smallest validation loss. The binary cross entropy was selected as the loss function, which measures the gap between predicted labels and true labels. Our model training process did not rely on a rigid learning rate. Instead, it leveraged the scheme of cyclical learning rates which could reduce the guesswork for selecting an optimal learning rate and demonstrates improved classification accuracy^21^.

The model training was performed on a computing server configured with a 24-core, 3.8GHz CPU, 128 GB DDR4 RAM, and a GPU with 10496 CUDA cores and 24GB GDDR6X graphics memory. Key software and packages used to train the deep learning models included fast.ai (ver.1.0.61), PyTorch (ver.1.11.0), and Python (ver.3.10.7).

### Performance evaluation and model interpretability

Outputs of machine learning models after the sigmoid activation function were probabilities that span continuously in the range between zero and one. It required a preset probability threshold to generate predicted labels, which were compared with true labels to obtain true positive (TP), false positive (FP), true negative (TN), and false negative (FN). These were the building blocks to calculate traditional performance metrics, such as accuracy, sensitivity, specificity, precision, and F1 scores. We first used the area under the receiver operating characteristic curve (AUROC) as the main metric to compare the performance of different models, because it is threshold agnostic and can serve as an aggregated metric to evaluate the overall performance of machine learning models. In addition, we provided the five traditional metrics mentioned above by using the default probability threshold of 0.5 to further demonstrate the model performance.

We delved into the interpretability of the proposed Multimodal model by calculating the SHapley Additive exPlanations (SHAP) values^22^. SHAP is a method based on cooperative game theory that can gauge the importance of each feature contributing to the model output. We compared the maximal feature importance from each feature category to elucidate their levels of contribution to the prediction of MI. Furthermore, feature importances across ECG tracings were evaluated to reveal regions of ECG beats to which the model paid heavy attention to.

### Experiment design

We designed two experiments to investigate two separate but important factors entrained in the development of machine learning models for predicting myocardial infarction. Experiment 1 concerns the duration of ECG waveforms entering model training. Clinical standard 12-lead ECGs are typically presented in a 3 by 4 matrix of 2.5s waveforms from each lead, sometimes accompanied by 10-second rhythmic strips stacked at the bottom. Clinicians evaluate the 2.5s of ECG waveforms for decision-making, despite digital recordings of full 10-second waveforms from all ECG leads are ubiquitously captured in modern ECG devices. In Experiment 1, we compared the prediction performance of ECG-only models trained from ECG samples of 2.5s and 10s, respectively. Such comparison helps answer the question of whether the ECG-only model can benefit from the additional temporal ECG waveforms to achieve improved performance.

There is rich digital information routinely captured during intitial evaluation of MI, in addition to the standard ECG. Such information potentially provides a more complete picture about a patient’s condition than the ECG alone. In Experiment 2, we added patient demographic information (i.e., age and sex) as a first step into the model design to obtain the Multimodal model and compare its prediction performance with the ECG-only model. The comparison outcome will elucidate whether leveraging additional clinical variables existing in the early clinical pipeline can help elevate the prediction power of machine learning models for detecting MI.

Given the small sample size of the model performance, the nonparametric two-sided Wilcoxon Rank Sum tests are adopted to compare the prediction performance (using AUROC) of different models in these experiments. The significance level is set at 0.05, which is adjusted with Bonferroni correction for multiple comparisons.

## Results

Table 1 shows the performance achieved by the ECG-only model that was trained with ECG waveforms of 2.5s. The mean AUROC achieved by the model was 89.8% which is averaged across 9 cross-validation folds. It also shows the performance is stable from models obtained from different CV folds with a small standard deviation (SD) of 0.5%. When setting the probability threshold to 0.5, the model obtains an overall accuracy of 84.7% (SD: 0.7%), and a sensitivity of 74.5% (SD: 2.4%) while offering a competitive specificity of 88% (SD: 1.3%). Given the imbalance numbers of samples across the MI and non-MI conditions (inter-class ratio: 1-3), it is essential to also include metrics resilient to class imbalance when evaluating the model performance. The model achieves a moderate positive predictive value (i.e., precision) of 67.7% (SD: 2%) and an F1 score (that is the harmonic mean of precision and recall) of 70.9% (SD: 1.1%).

**Table 1.** Prediction performance of the ECG-only model trained with 2.5s ECG.

| CV Fold | AUROC | Accuracy | Sensitivity | Specificity | Precision | F1 |
| --- | --- | --- | --- | --- | --- | --- |
| 1 | 0.893 | 0.840 | 0.736 | 0.875 | 0.663 | 0.698 |
| 2 | 0.895 | 0.839 | 0.781 | 0.859 | 0.650 | 0.709 |
| 3 | 0.899 | 0.841 | 0.756 | 0.869 | 0.659 | 0.704 |
| 4 | 0.905 | 0.852 | 0.765 | 0.881 | 0.683 | 0.722 |
| 5 | 0.893 | 0.847 | 0.696 | 0.898 | 0.696 | 0.696 |
| 6 | 0.893 | 0.843 | 0.738 | 0.879 | 0.671 | 0.703 |
| 7 | 0.902 | 0.846 | 0.736 | 0.883 | 0.678 | 0.706 |
| 8 | 0.899 | 0.849 | 0.756 | 0.880 | 0.679 | 0.715 |
| 9 | 0.902 | 0.861 | 0.743 | 0.900 | 0.714 | 0.728 |
| Mean/SD | 0.898/0.005 | 0.847/0.007 | 0.745/0.024 | 0.880/0.013 | 0.677/0.020 | 0.709/0.011 |

Abbreviations: CV = cross-validation; SD = standard deviation; AUROC = area under the receiver operating characteristic curve.

Table 2 presents the performance achieved by the ECG-only model that was trained with 10s ECG waveforms. When compared to the 2.5s model as designed in Experiment 1, the 10s model improves the overall AUROC and accuracy, reaching 91.5% (SD: 0.4%) and 87.1% (SD: 0.4%), respectively. With a probability threshold of 0.5, the 10s model also offers improved specificity at 92.9% (SD: 0.5%) over the 2.5s model but at the cost of a decrease in sensitivity at 69.9% (SD: 1.5%). There is also a great improvement in the precision and the F1 score, which are 76.8% (SD: 1.1%) and 73.2% (SD: 0.9%) from the 10s model. The two-sided Wilcoxon Rank Sum test shows that there is a statistical significance in the AUROC achieved by both models (*p<0*.*001*), with the 10s model outperforming the 2.5s model by 1.7% (see first two bars in Fig. 3).

**Table 2.**
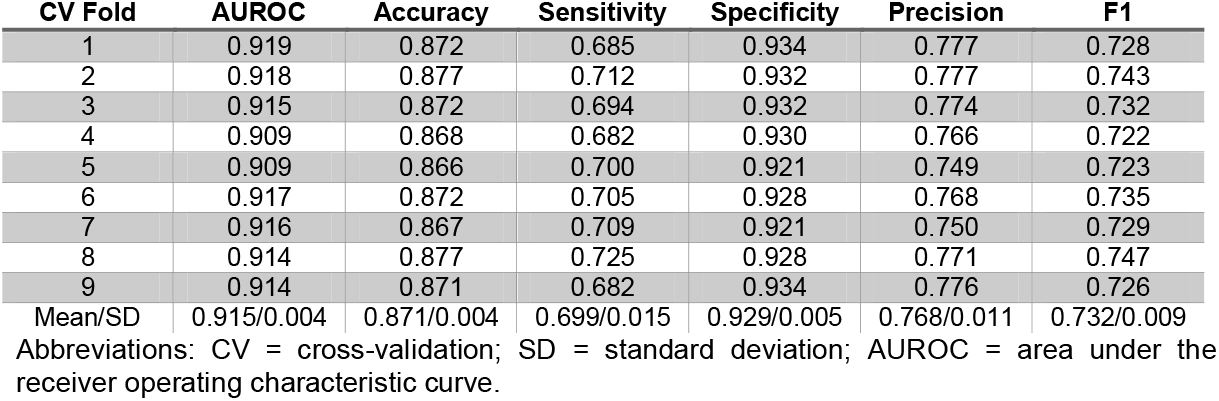
Prediction performance of the ECG-only model trained with 10s ECG.

**Figure 3.**
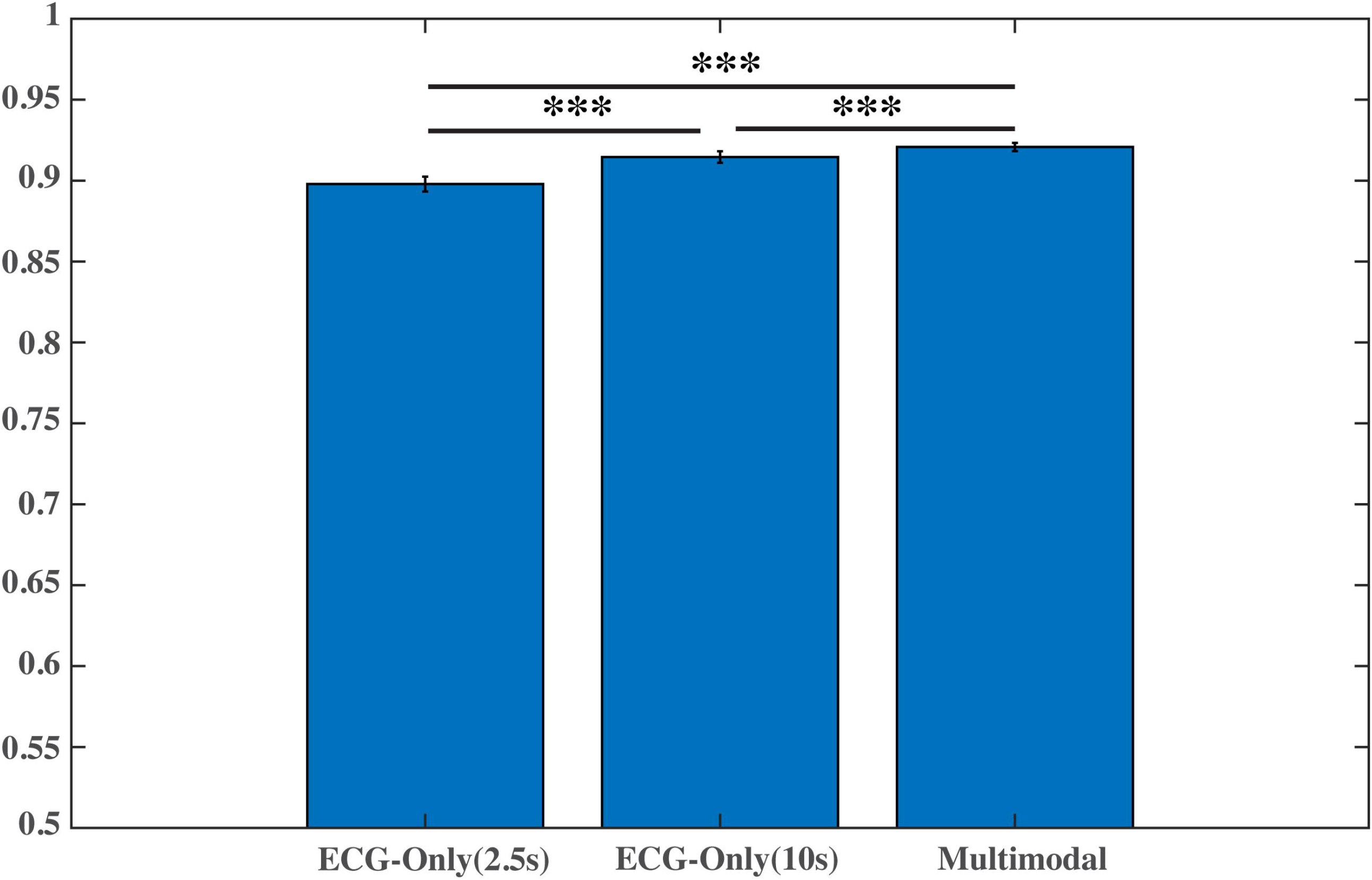
Performance comparison between ECG-only models of different waveform durations and the Multimodal model (*** indicates *p<0*.*001*).

Table 3 shows the performance acquired with the Multimodal model that integrates 10s ECG and patient demographic information as model inputs. Results show that the Multimodal model continues to improve performance metrics that reflect aggregated model performance, with increased AUROC (mean/SD: 92.1%/0.3%), accuracy (mean/SD: 87.4%/0.3%), precision (mean/SD: 77.9%%/1.2%), and F1 score (mean/SD: 73.5%/0.8%), compared to ECG-only models. Notably, the Multimodal model achieved comparable sensitivity (mean/SD: 69.7%/1.6%) to the 10s ECG-only model, while offering improved specificity at 93.4% (SD: 0.6%). The two-sided Wilcoxon Rank Sum confirms the statistical significance in the AUROC achieved by the two models (*p<0*.*001*) (see second and third bars in Fig. 3).

**Table 3.** Prediction performance of the Multimodal model trained with a combination of 10s ECG and patient demographic information.

| CV Fold | AUROC | Accuracy | Sensitivity | Specificity | Precision | F1 |
| --- | --- | --- | --- | --- | --- | --- |
| 1 | 0.923 | 0.873 | 0.673 | 0.941 | 0.791 | 0.727 |
| 2 | 0.919 | 0.875 | 0.684 | 0.939 | 0.789 | 0.733 |
| 3 | 0.925 | 0.879 | 0.723 | 0.932 | 0.780 | 0.750 |
| 4 | 0.917 | 0.871 | 0.694 | 0.930 | 0.770 | 0.730 |
| 5 | 0.918 | 0.872 | 0.682 | 0.935 | 0.779 | 0.727 |
| 6 | 0.922 | 0.874 | 0.714 | 0.927 | 0.767 | 0.74 |
| 7 | 0.921 | 0.875 | 0.691 | 0.937 | 0.786 | 0.735 |
| 8 | 0.920 | 0.870 | 0.707 | 0.924 | 0.758 | 0.732 |
| 9 | 0.922 | 0.879 | 0.703 | 0.938 | 0.791 | 0.744 |
| Mean/SD | 0.921/0.003 | 0.874/0.003 | 0.697/0.016 | 0.934/0.006 | 0.779/0.012 | 0.735/0.008 |

Abbreviations: CV = cross-validation; SD = standard deviation; AUROC = area under the receiver operating characteristic curve.

Figure 4 presents the feature importance of different types of features used by the model, and feature importance along the ECG tracing. The top panel reveals that the most important feature contributing to the model performance is ECG, followed by se and age in MI samples. The ranking stays true in control samples but with reduced importance for all feature categories. The bottom panel presents feature importance across the first 5 seconds of ECG in a MI sample on Lead aVF. There is a clear ST-segment elevation presented in the ECG. The model can be seen paying heavy attention to several ECG landmarks, including the J point, ST segments, and the QRS complex.

**Figure 4.**
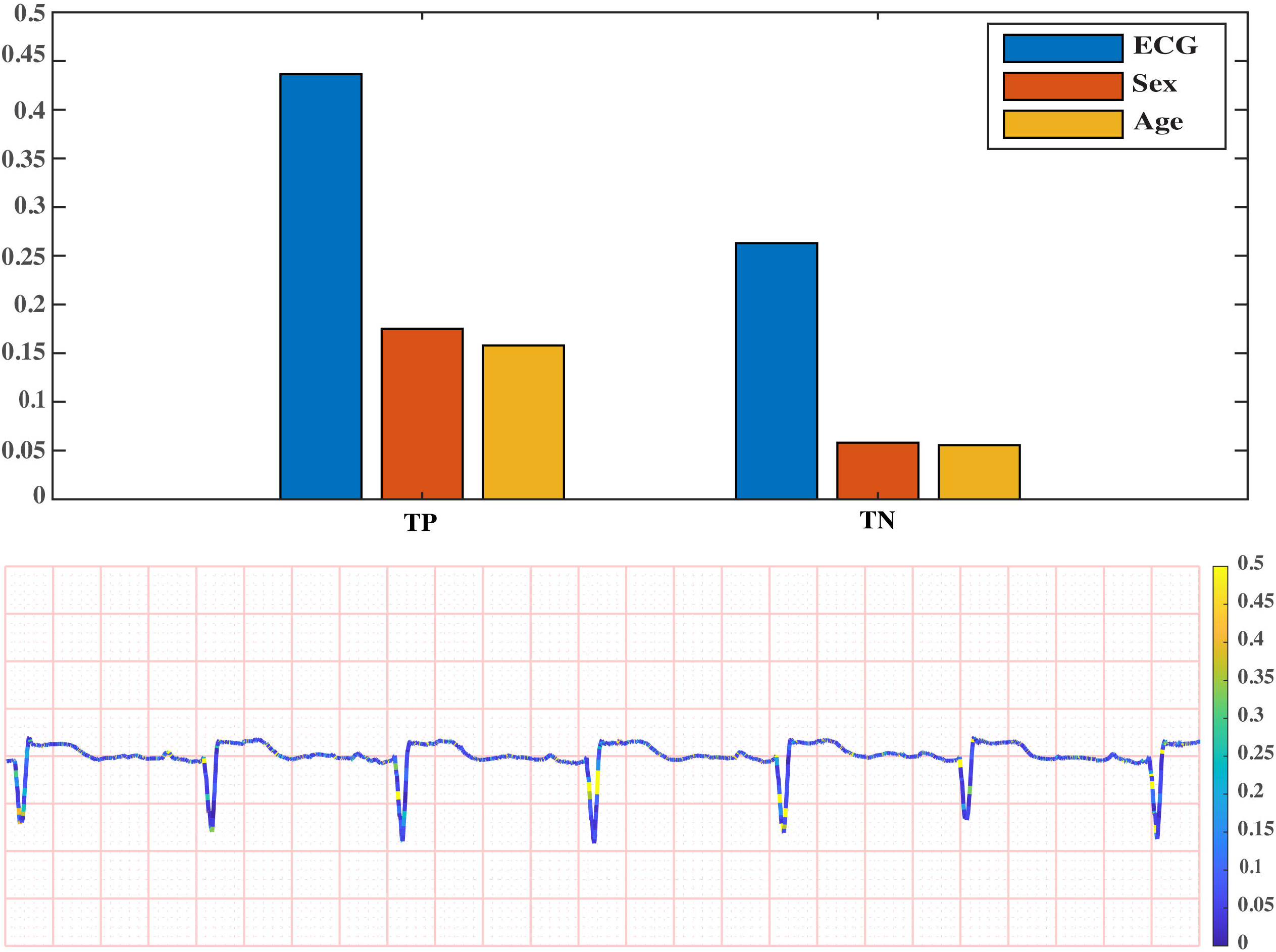
Feature importance in the Multimodal model. **Top panel**: comparison of feature importance among three feature categories. **Bottom panel**: feature importance in ECG tracings. The ECG is from a MI sample and the plot contains the first 5 seconds on Lead aVF.

## Discussion

The key findings of this study are that the multimodal information helps improve the predictivity of machine learning models for early identification of myocardial infarction and that the data-driven approach can leverage the extra length of ECG waveforms to make a better prediction. The study addresses a critical unmet need for early and accurate identification of acute myocardial infarction during the initial evaluation phase. Recognizing the underlying collusion of coronary arteries early in time is associated with improved patient outcomes, as it helps reduce door-to-balloon time by, 1. optimizing EMS routes by sending MI patients directly to the nearest hospital with PCI-capability; 2. alerting the Cath Lab Team in advance allowing extra time for the preparation of revascularization therapies. Reliable prediction can also reduce false activation of the cardiac catheterization lab which saves the hospital operation cost and reserves limited clinical resources for patients in urgent need. With that in mind, our study purposefully constrained the types of data entered into model training to those commonly available during the initial care stage for MI. In addition, we dichotomized ECG labels into the MI class vs. all other conditions, including both normal ECGs and chronic conditions that might confound the initial diagnosis of MI, such as LBBB and LVH. Such dichotomization matches the clinical scenario for early screening MI. Under this direction, we investigated two important factors embroiled in the decision process of developing MI prediction models using machine learning, i.e., data length and data types, which concern how much and what information to enter model training, respectively.

Standard 12-lead ECGs are conventionally represented in the 2.5s fashion as a convenient layout to provide an overview of information from all leads on a single screen. Healthcare providers are trained to make clinical inferences based on the standard 2.5s layout. However, modern ECG machines store full 10s of ECG tracings, which are not fully utilized in current clinical practice. The added duration in ECG may offer extra electrophysiological dynamics of hearts that help decypher the ongoing development of myocardial ischemia, given that acute MI is a rapidly developing condition. Machine learning by nature is a data-driven approach that can simultaneously sift through information well-beyond human compacity, therefore may capitalize on the additional information. This is demonstrated by results from Experiment 1 (see Fig. 3) that a significantly improved prediction performance can be obtained when using 10s ECG waveforms to train the same deep neural network as the 2.5s model.

Although standard 12-lead ECG remains the single most important clinical tool in the initial evaluation of myocardial infarction, mounting evidence suggests there is value to integration of other information during the risk assessment process^16^. For example, studies show there is a sex difference in electrocardiographic manifestations of acute myocardial ischemia, with women presenting a lower ST elevation^23^. It also shows that there is a decrease in ST elevation indicative of acute myocardial ischemia with increasing age^16^. Therefore, both age and sex have been incorporated into the latest universal definition of myocardial infarction^16^. In addition, patients with a previous diagnosis or at high risk of coronary artery disease may have altered ECG presentations^24^. The inclusion of such information is therefore recommended by the international guideline to enhance the specificity of ECG findings^16^. In the present study, we designed a feature-infusion deep learning model that learns multimodal information including the patient demographics and physiologic information from ECG and compared the obtained performance with the ECG-only model. Results from Experiment 2 (see Fig. 3) show a significantly improved performance by the Multimodal model, demonstrating that the added patient demographics contribute to the model learning process by providing age and sex-adjusted discriminative features for the prediction task.

Past studies have recognized the promise of machine learning in predicting myocardial infarction using ECG^10-14, 25^. However, most of the existing studies formulated the classification problem (i.e., MI vs. Normal) in facilitation of identifying ECG features reflecting underlying ischemia at the expense of derailing from practical clinical applications, where both normal and other non-MI, even MI-confounding conditions exist besides MI. Our study is a big proponent of bearing this clinical need from the start, through the dichotomization of labels into MI and non-MI classes. Machine learning algorithms adopted in existing studies can be separated into shallow machine learning and deep learning. Our study followed the latter choice as we took advantage of the large public ECG dataset that provides an ample number of samples to train a sophisticated deep neural network (i.e., xResNet)^15^. Comparison of performance across these studies is challenging due to the wide variety of experimental designs and performance metrics being measured. Here we selected accuracy as the metric to provide a rough sense of where our model performance stands comparing with others, as it is commonly reported across studies. Despite the inclusion of confounding conditions in the control class that challenges the classification task in our study, the accuracy of 87.4% achieved by the Multimodal model is on par with other studies, which reported accuracies in the range of 82.5%∼93.7%^10-14, 25^.

The present study aims to set the ground for early detection of myocardial infarction using machine learning that is suitable in clinical settings. This is achieved by dividing sample labels with consideration of downstream clinical applications and only utilizing data available during the early evaluation phase of MI for model development. There are several limitations in the study that will need future works to address. The Multimodal model only included patient demographics as the additional data for model development, while other information, such as patient symptoms and past diagnoses, is also routinely collected during the initial care stage and has great potential to further elevate the prediction performance. Our study is limited in that the dataset doesn’t contain the above information, but the unique network design of the proposed Multimodal model makes it readily expandable to other data modalities by simply adding new embedding modules. Therefore, a future study is warranted to investigate the value of those other data modalities. In addition, the dataset adopted in the study was collected from multiple centers and thousands of patients, which intrinsically offers a sample distribution with a strong estimation of the overall population. Nonetheless, the obtained model still requires extensive external validation to establish its generalizability, which is the key to successful model deployment.

## Data Availability

All data produced are available online at the PhysioNet website.
https://physionet.org/content/ptb-xl/1.0.2/

## References

1. Al-Zaiti SS, Alghwiri AA, Hu X, Clermont G, Peace A, Macfarlane P, Bond R. A clinician’s guide to understanding and critically appraising machine learning studies: a checklist for Ruling Out Bias Using Standard Tools in Machine Learning (ROBUST-ML). European Heart Journal - Digital Health. 2022;3(2):125–40.

2. Al-Zaiti S, Macleod R, Dam PV, Smith SW, Birnbaum Y. Emerging ECG methods for acute coronary syndrome detection: Recommendations & future opportunities. J Electrocardiol. 2022;74:65–72. Epub 2022/08/27. PubMed PMID: 36027675.

3. Farquharson B, Abhyankar P, Smith K, Dombrowski SU, Treweek S, Dougall N, Williams B, Johnston M. Reducing delay in patients with acute coronary syndrome and other time-critical conditions: a systematic review to identify the behaviour change techniques associated with effective interventions. Open heart. 2019;6(1):e000975. Epub 2019/04/19. PubMed PMID: 30997136; PMCID: PMC6443141.

4. Goldman L, Kirtane AJ. Triage of patients with acute chest pain and possible cardiac ischemia: the elusive search for diagnostic perfection. Ann Intern Med. 2003;139(12):987–95. Epub 2003/12/18. PubMed PMID: 14678918.

5. Fesmire FM, Percy RF, Bardoner JB, Wharton DR, Calhoun FB. Usefulness of automated serial 12-lead ECG monitoring during the initial emergency department evaluation of patients with chest pain. Annals of Emergency Medicine. 1998;31(1):3–11. Epub 1998/01/23. PubMed PMID: 9437335.

6. Kudenchuk PJ, Maynard C, Cobb LA, Wirkus M, Martin JS, Kennedy JW, Weaver WD. Utility of the prehospital electrocardiogram in diagnosing acute coronary syndromes: the Myocardial Infarction Triage and Intervention (MITI) Project. Journal of the American College of Cardiology. 1998;32(1):17–27. Epub 1998/07/21. PubMed PMID: 9669244.

7. Forberg JL, Green M, Bjork J, Ohlsson M, Edenbrandt L, Ohlin H, Ekelund U. In search of the best method to predict acute coronary syndrome using only the electrocardiogram from the emergency department. Journal of Electrocardiology. 2009;42(1):58–63. Epub 2008/09/23. PubMed PMID: 18804783.

8. Rouan GW, Lee TH, Cook EF, Brand DA, Weisberg MC, Goldman L. Clinical characteristics and outcome of acute myocardial infarction in patients with initially normal or nonspecific electrocardiograms (a report from the Multicenter Chest Pain Study). American Journal of Cardiology. 1989;64(18):1087–92. Epub 1989/11/15. PubMed PMID: 2683709.

9. Chantad D, Krittayaphong R, Komoltri C. Derived 12-lead electrocardiogram in the assessment of ST-segment deviation and cardiac rhythm. Journal of Electrocardiology. 2006;39(1):7–12. Epub 2006/01/03. PubMed PMID: 16387043.

10. Wang Z, Qian L, Han C, Shi L. Application of multi-feature fusion and random forests to the automated detection of myocardial infarction. Cognitive Systems Research. 2020;59:15–26.

11. Chang P-C, Lin J-J, Hsieh J-C, Weng J. Myocardial infarction classification with multi-lead ECG using hidden Markov models and Gaussian mixture models. Applied Soft Computing. 2012;12(10):3165–75.

12. Sharma LN, Tripathy RK, Dandapat S. Multiscale Energy and Eigenspace Approach to Detection and Localization of Myocardial Infarction. IEEE Transactions on Biomedical Engineering. 2015;62(7):1827–37.

13. Reasat T, Shahnaz C, editors. Detection of inferior myocardial infarction using shallow convolutional neural networks. 2017 IEEE Region 10 Humanitarian Technology Conference (R10-HTC); 2017 21-23 Dec. 2017.

14. Strodthoff N, Wagner P, Schaeffter T, Samek W. Deep Learning for ECG Analysis: Benchmarks and Insights from PTB-XL. IEEE journal of biomedical and health informatics. 2021;25(5):1519–28. Epub 20210511. PubMed PMID: 32903191.

15. Wagner P, Strodthoff N, Bousseljot RD, Kreiseler D, Lunze FI, Samek W, Schaeffter T. PTB-XL, a large publicly available electrocardiography dataset. Sci Data. 2020;7(1):154. Epub 20200525. PubMed PMID: 32451379; PMCID: PMC7248071.

16. Thygesen K, Alpert JS, Jaffe AS, Chaitman BR, Bax JJ, Morrow DA, White HD, Executive Group on behalf of the Joint European Society of Cardiology /American College of Cardiology /American Heart Association /World Heart Federation Task Force for the Universal Definition of Myocardial I. Fourth Universal Definition of Myocardial Infarction (2018). Circulation. 2018;138(20):e618-e51. PubMed PMID: 30571511.

17. Goldberger AL, Amaral LA, Glass L, Hausdorff JM, Ivanov PC, Mark RG, Mietus JE, Moody GB, Peng CK, Stanley HE. PhysioBank, PhysioToolkit, and PhysioNet: components of a new research resource for complex physiologic signals. Circulation. 2000;101(23):E215-20. PubMed PMID: 10851218.

18. International Organization for Standardization. ISO 11073 91064:2009, Health Informatics—Standard Communication Protocol—Part 91064: Computer-Assisted Electrocardiography; ISO: Geneva, Switzerland, 2009; pp. 1–159.

19. He T, Zhang Z, Zhang H, Zhang Z, Xie J, Li M. Bag of Tricks for Image Classification with Convolutional Neural Networks 2018 December 01, 2018:[1812.01187 p.]. Available from: https://ui.adsabs.harvard.edu/abs/2018arXiv181201187H.

20. He K, Zhang X, Ren S, Sun J. Deep Residual Learning for Image Recognition 2015 December 01, 2015:[1512.03385 p.]. Available from: https://ui.adsabs.harvard.edu/abs/2015arXiv151203385H.

21. Smith LN. Cyclical Learning Rates for Training Neural Networks 2015 June 01, 2015:[1506.01186 p.]. Available from: https://ui.adsabs.harvard.edu/abs/2015arXiv150601186S.

22. Lundberg S, Lee S-I. A Unified Approach to Interpreting Model Predictions 2017 May 01, 2017:[1705.07874 p.]. Available from: https://ui.adsabs.harvard.edu/abs/2017arXiv170507874L.

23. Europa WRf. Ischemic Heart Disease Registers: Report of the Fifth Working Group (including a Second Revision of the Operating Protocol), Convened by the Regional Office for Europe of the World Health Organization ; Copenhagen, 26-29 April 1971 : [Fifth Working Group on Ischaemic Heart Disease Registers] 1971.

24. Delewi R, Ijff G, van de Hoef TP, Hirsch A, Robbers LF, Nijveldt R, van der Laan AM, van der Vleuten PA, Lucas C, Tijssen JG, van Rossum AC, Zijlstra F, Piek JJ. Pathological Q waves in myocardial infarction in patients treated by primary PCI. JACC Cardiovasc Imaging. 2013;6(3):324–31. Epub 20130220. PubMed PMID: 23433932.

25. Dohare AK, Kumar V, Kumar R. Detection of myocardial infarction in 12 lead ECG using support vector machine. Applied Soft Computing. 2018;64:138–47.

